# Prenatal glycolysis restoration can rescue myocardial hypoplasia caused by platelet isoform of phosphofructokinase 1(PFKP) deficiency

**DOI:** 10.1101/2024.01.07.24300871

**Authors:** Siyao Zhang, Hairui Sun, Xiaoyan Hao, Xu Zhi, Ruimin Liu, Tong Yi, Ye Zhang, Xiaoyan Gu, Jiancheng Han, Xiaowei Li, Jiaqi Fan, LiYing Yan, Hankui Liu, Feng Lan, Hongjia Zhang, Jie Qiao, Yihua He

**Author notes:** Contributed equally (Siyao Zhang, Hairui Sun, Xiaoyan Hao, Xu Zhi, Ruimin Liu). Correspondences: Feng Lan; Hongjia Zhang; Jie Qiao (.); Yihua He. Lead Contact: Yihua He.

## Abstract

**Background:** Congenital myocardial hypoplasia affects heart function in congenital heart diseases, but its causes and mechanisms are unclear. **Methods:** Fetuses with myocardial hypoplasia were examined using echocardiography imaging and genetic testing. The identified pathogenic genes were genetically targeted to validate mechanistic findings. We used stem cells and transgenic mice to understand molecular mechanisms and applied Preimplantation Genetic Testing for monogenic defects to obtain healthy offspring. In addition, 1,300 genetic sequencing records were screened to understand the prevalence of the disease and deepen our understanding of myocardial hypoplasia treatment.

**Results:** This is the first study to link PFKP pathogenic variant to human myocardial hypoplasia. We found that PFKP deficiency decreased embryonic heart glycolysis, resulting in a thinning myocardial wall and impaired cardiac function, attributable to a decline in cardiomyocyte proliferation. The intrauterine supplement with Fructose 1,6-bisphosphate, a direct product of PFKP catalysis, can rescue the main myocardial phenotype of fetal mice. Assisted reproductive technology was used to prevent PFKP pathogenic variant transmission to offspring. Finally, one of the family lines (family 1) obtained a healthy offspring with a normal heart. **Conclusions** PFKP plays a key role in regulating glycolysis during embryonic cardiac development. Addressing glycolytic defects is crucial for myocardial hypoplasia. We provide new insights that have implications for genetic interventions, prenatal screening, and targeted intervention strategies.

## Introduction

The congenital heart diseases (CHDs) have a prevalence of 8-12‰, rendering it the most prevalent birth defect worldwide. Additionally, it ranks as the leading cause of neonatal mortality^1, 2^. The causes of CHDs are complex and characterized by poorly elucidated factors and mechanisms, thereby posing significant challenges in formulating effective preventive and therapeutic strategies. Historically, research efforts have predominantly concentrated on postnatal diagnostics, mainly attributable to the constraints of available diagnostic modalities in capturing nuanced prenatal cardiac disease phenotypes with the requisite precision^3^. Nevertheless, a certain proportion of complex CHDs result in intrauterine and pre-hospital deaths, contributing to an incomplete understanding of the disease spectrum^4^.

Some CHD are accompanied by congenital myocardial hypoplasia^5, 6^, which includes conditions like congenital ventricular aneurysms^7, 8^, cardiac dverticula^7, 8^, primary cardiomyopathies^9^ and others^10, 11^. CHDs, whether characterized by concomitant structural anomalies or not, frequently manifest myocardial hypoplasia^12, 13^. Accompanying the advancement of technology, considerable advancements have been accomplished by developmental biologists in elucidating the genetic mechanisms of early cardiac development^11, 14^. Embryonic cardiomyocytes exhibit a remarkable proliferative capacity during fetal development, enabling them to proliferate efficiently and form a functional heart^15-17^. Cardiomyocyte proliferation is responsible for cardiac expansion during mammalian embryonic development. After birth, mammalian cardiomyocytes transition from proliferative growth to hypertrophic growth, marked by reduced cell cycle activity^18^. This transition represents a pivotal moment, precipitating substantial structural transformations and arguably distinct gene expression strategies between fetal cardiac development and postnatal cardiac maturation^19^. Despite this commonality, the genetic mechanisms governing embryonic development remain elusive. The delineation of shared pathological mechanisms and genetic regulatory pathways holds profound significance in the context of preventing and intervening in congenital heart malformations.

Our investigation, conducted within the distinctive temporal window of prenatal cardiac embryonic development via genomic sequencing and rodent model investigations, has delineated novel clinical phenotypes and functional deficiencies associated with the *PFKP* gene, which broadens the disease spectrum of CHDs. More importantly, we revealed the pivotal role and mechanistic underpinnings of *PFKP* defects during various stages of embryonic development. This influence is manifested through the modulation of anaerobic glycolytic energy metabolism, downstream metabolites, and the consequential reduction in myocardial proliferative capacity, ultimately culminating in compromised cardiac development. Furthermore, our study has successfully ameliorated the phenotypic manifestations of impaired cardiac development through the strategic supplementation of downstream metabolites. Consequently, these findings offer a compelling foundation for genetic interventions and exhibit the potential to identify effective intrauterine intervention targets by ameliorating associated phenotypes.

## Methods

### Study subjects

Thorough clinical evaluations of affected fetuses and adults associated with Family 1 and Family 2 members were performed, employing diagnostic modalities including echocardiography, cardiovascular magnetic resonance (CMR), and electrocardiograms. The DNA of affected individuals and control individuals from both families underwent rigorous whole-genome sequencing (WGS) analysis. For more details, please refer to Appendix 1. Informed written consent was obtained from all study participants or their legal guardians, and the study was approved by the Ethics Committee of Beijing Anzhen Hospital of Capital Medical University. The authors ensure the accuracy and completeness of the data in this report.

### Mouse model

The establishment of a mutation knock-in mouse model harboring the *Pfkp* R754W pathogenic site, analogous to the identified mutation in the patient, was carried out concurrently with the development of a myocardial-specific *Pfkp* conditional knockout (cKO) mouse model. A comprehensive description of mouse model generation is provided in Supplementary Appendix 1.

### Human embryonic stem cells (hESC) and human induced pluripotent stem cells (hiPS)

The *PFKP* gene knockout in hESC was achieved utilizing Clustered Regularly Interspaced Short Palindromic Repeats technology. The resulting modified human pluripotent stem cells (hPSCs) were then guided through specific media to induce differentiation into cardiomyocytes, facilitating subsequent cell studies.

Peripheral blood mononuclear cells (PBMC) were extracted from both the patient (III-4) and unaffected family members (III7). Subsequently, these PBMCs were reprogrammed into hiPS, wherein the *PFKP* R755W mutation was corrected through homologous recombination. This correction yielded hiPS (*PFKP* Correct/+) possessing the identical genetic background as the patient but devoid of the PFKP R755W mutation.

A comprehensive account of the methodology employed in generating human pluripotent stem cell-derived cardiomyocytes (hPSC-CMs) and other pertinent laboratory research endeavors is provided in Supplementary Appendix 1.

### Assisted Reproductive Technology

Genetic testing using the Mutated Allele Revealed by Sequencing with Aneuploidy and Linkage Analyses (MARSALA)^20^ technology was conducted on embryos obtained from Family 1. An embryo devoid of the *PFKP* R755W variant was meticulously selected and subsequently transplanted into the maternal uterus. Following successful implantation, a comprehensive regimen of prenatal examinations and fetal echocardiography was performed, with ongoing postnatal follow-up extending to eighty days after birth.

### Statistical Analysis

Descriptive statistics for continuous variables are shown as the means ± standard deviation, while categorical variables are summarized with subject counts and percentages. Unpaired two-tailed Student’s t-test were used for statistical comparison of two groups. For non-normal data, non-parametric Mann-Whitney test was used. Statistical analysis was performed using Prism 8.0 (GraphPad Software). A value of P < 0.05 was considered statistically significant.

## Results

### Clinical presentation and identification of *PFKP* pathogenic variant

We conducted an in-depth examination of the family where the proband was discovered. Fetal echocardiography, revealed myocardial hypoplasia in the developing fetus (Fig. 1a, IV-2), conceived by unrelated parents (Fig. 1a). This fetus has secondary cardiac dysfunction, as evidenced by a Cardiovascular Profile Score (CVPS) of 7 points. The father (Fig. 1a, III-2) exhibits a similar clinical phenotype, with a 40% ejection fraction and heart failure (Fig. 1a). The echocardiography of the fetus revealed myocardial thinning in the ventricular septum and the presence of a congenital ventricular aneurysms in the middle to an apical segment of the left ventricular free wall (Fig.1b). Considering the identified cardiac anomalies in the current pregnancy, along with the parents’ history of a prior adverse pregnancy resulting in fetal demise, an unfavorable prognosis was indicated. Consequently, the parents opted for induced labor.

**Figure 1.**
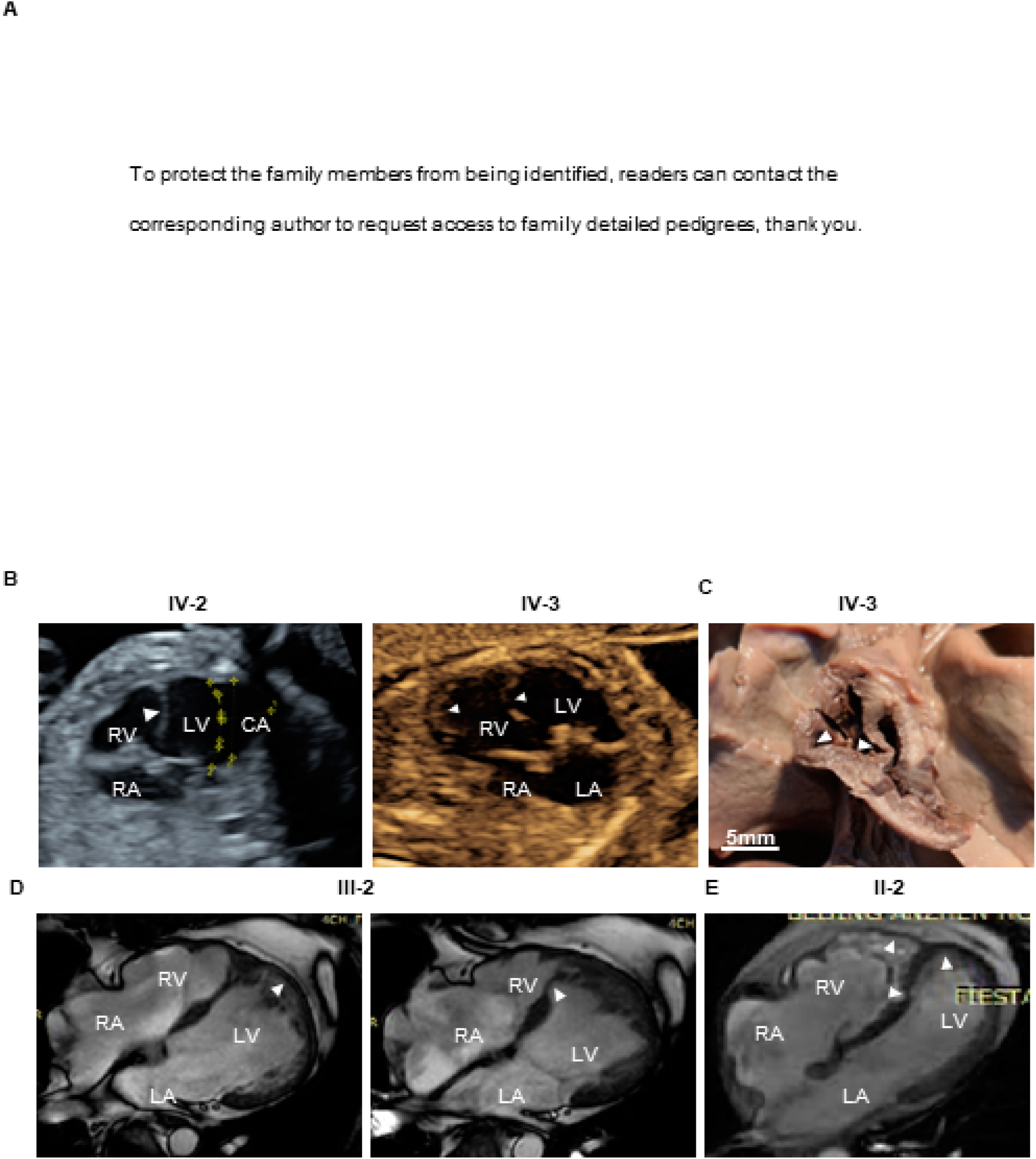
Identification of *PFKP* R755W variant in patients with myocardial hypoplasia. **Panel A** shows the pedigree of Family 1 with myocardial hypoplasia. Family members for whom WGS were obtained are indicated as Wt/Mut or Wt/Wt. Squares represent male family members, circles represent female family members, and triangles represent the unborn fetus. Solid grey shapes indicate affected persons. Question marks indicate individuals whose clinical status is uncertain. Slashed symbols indicate deceased individuals. Arrows indicate probands. Wt/Mut indicates *PFKP* R755W heterozygous variation; Wt/Wt indicates R755 wild type. **Panel B** shows the echocardiography of the affected fetus (IV-2 and IV-3). Patient IV-2 showed thinning of the ventricular septum, the myocardium in the middle and apical segments of the left ventricular free wall, and CA (The arrow points to the bulging ventricular septum). IV-3 showed thinning of the left and right ventricular walls and the ventricular septum, and the middle segment of the right ventricular free wall myocardial bulge (The arrow indicates the bulge of the ventricular septum and the right ventricular myocardium). **Panel C** shows a section of the postmortem heart of the IV-3 patient, in which the septal myocardium of the left and right ventricular walls are thinner than average. The ventricular septal myocardium and right ventricular wall are more prominent (arrowhead). **Panels D and E** show cardiovascular magnetic resonance (CMR) images of the affected adults (III-2 and II-2). The left image in Panel D shows a blunt left ventricular apex(indicated by an arrowhead) and right ventricular enlargement in patient III-2. The right image in Panel D shows thinning of the interventricular septum with bulges toward the right ventricle side (indicated by an arrowhead) in patient III-2. Panel E exhibits thinning of the right ventricular myocardium (indicated by the left arrowhead) and the ventricular septum, bulging toward the left ventricular side (indicated by the right arrowhead) and a blunt left ventricular (indicated by the up arrowhead). CA: congenital ventricular aneurysms; LA: left atrium; LV: left ventricle; RA: right atrium; RV: right ventricle; WGS: whole genome sequencing.

Subsequently, in the following pregnancy of the same mother (Fig. 1a, IV-3), fetal echocardiography revealed a cardiac phenotype strikingly consistent with the previous one (Fig. 1a, IV-2), and once again, clinical manifestations of thinning of the left and right ventricular walls and septal myocardium, along with cardiac insufficiency resulting in a CVPS of 6 points. Informed of these findings, the family decided to terminate the pregnancy for a second time and proceed with a postmortem examination, which confirmed the prenatal diagnosis (Fig.1c). Upon an exhaustive review of the family’s medical history, a striking pattern emerged, demonstrating that this condition had afflicted members across three consecutive generations. Importantly, most affected family members exhibit concurrent structural cardiac abnormalities, such as atrial or ventricular septal defects, alongside right bundle branch block (Table 1). The consistent phenotype observed within this familial lineage aligns with an autosomal dominant mode of inheritance (Fig.1a).

**Table1.** Symptoms of patients with myocardial hypoplasia (readers contact the corresponding author to request access to these data)

WGS was performed in the family to identify the genetic etiology. Initial investigations into genes associated with CHD failed to yield any positive results. However, a comprehensive genome-wide linkage analysis uncovered a co-segregating region spanning 5.8 Mb on chromosome 10 (p15.3-p15.1), displaying a logarithm of odds ratio (LOD) score of 2.4, surpassing the anticipated LOD threshold of 2.3. Further analysis of rare, damaging variants co-segregated with cardiac phenotypes within the family identified a heterozygous missense variant of *PFKP* gene (NM_002627.4:c.2263C>T:p.R755W). The *PFKP* R755W variant localizes within the co-segregation region and has a full likelihood Bayes factor (FLB) of 127.6, providing strong evidence in support of its pathogenicity. The *PFKP* R755 residue is highly conserved in vertebrates, yet R755W is absent from the Genome Aggregation Database. (http://gnomad-sg.org/).

Finally, structural and functional analyses of the PFKP protein were performed, revealing that the R755W variant was predicted to disrupt the protein’s configuration and functionality. To assess the impact of these patient-specific mutations on PFK1 enzymatic activity in HEK293T cells, we transfected PFKP wild type (WT) and PFKP mutant proteins by lentivirus and the expression of exogenous proteins was monitored. Interestingly, cells carrying the mutation displayed a marked 58% reduction in PFK1 enzymatic activity.

To provide a comprehensive understanding of PFKP’s expression patterns during mammalian heart development, gene expression data imported from Expression Atlas (https://www.ebi.ac.uk/gxa/home) was analyzed. Our results showed that PFKP is strongly expressed in fetal hearts, especially compared to other organs. Moreover, we observed high expression levels of PFKP in both human and mouse embryonic hearts. To better understand how PFKP changes during human cardiomyocyte development and to accommodate for species variations, we analyzed hiPS-CMs at different developmental stages. Notably, PFKP expression peaked in immature cardiomyocytes, specifically in 10-day and 15-day hiPS-CMs. These findings collectively indicate the pivotal role that PFKP likely plays in embryonic heart development.

### Mimicking Myocardial Hypoplasia Phenotype in Patients via a Pfkp^R754W/R754W^ Mice Model

To determine the phenotype of myocardial hypoplasia in affected families due to the *PFKP* R755W variants, we engineered a mouse model carrying the *PFKP* R755W equivalent variant (mouse *Pfkp* R754W). Subsequent analyses included confirming *Pfkp* mutant sequences and quantifying protein expression levels. *PFKP* mutation does not affect PFKP protein expression levels, but it did lead to a notable reduction in PFK1 enzyme activity.

Then, we performed histological analyses, the hearts of heterozygous, homozygous knock-in embryos, and WT embryos were examined, spanning the developmental period from E13.5 to E17.5, with a specific focus on heart development. Pfkp^R754W/+^ and Pfkp^R754W/R754W^ mice were viable and fertile. Upon a thorough examination of the left ventricular (LV) and right ventricular (RV) compact layer thickness, heterozygous Pfkp^R754W/+^ mice exhibited no overt phenotypic anomalies during the developmental stages spanning from E13.5 to E17.5. Conversely, Pfkp^R754W/R754W^ mice showed no significant changes in the LV and RV compact layer morphology at E13.5, but a significant thinning of the LV compact layer was observed at E15.5 and E17.5, while the thickness of the RV compact layer remained largely unaffected (Fig2a-2c). There was no significant difference in embryonic heart size between wild-type mice and mice carrying Pfkp^R754W/+^ or Pfkp^R754W/R754W^. These findings demonstrated that the Pfkp^R754W/R754W^ pathological outcomes were consistent with the patients’ clinical phenotype, affirming that the *PFKP* R755W mutation is the pathogenic variant underlying the clinical phenotype of myocardial hypoplasia.

**Figure 2.**
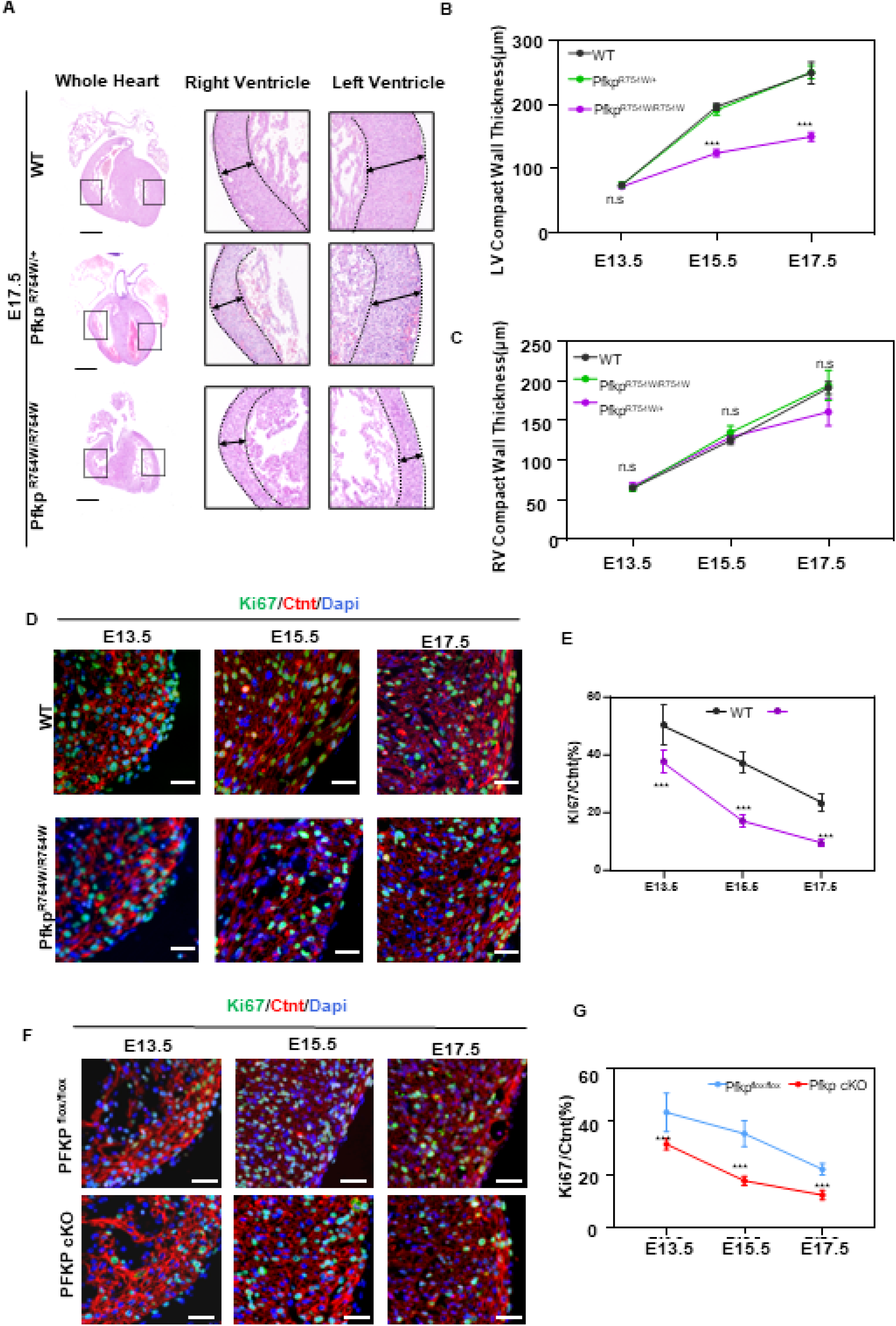
PFKP dysfunction results in decreased myocardial proliferation during the embryonic period. **Panel A** shows whole embryonic heart Hematoxylin-Eosin (HE) staining of representative Pfkp^R754W/+^, Pfkp^R754W/R754W^and WT mice at E17.5 with the coronal view, revealing that left ventricular walls of Pfkp^R754W/R754W^ mice are thinner than those of normal controls. The scale bar indicates 600μm. **Panel B and C** show quantification of left and right ventricular wall thickness between the Pfkp^R754W/+^, Pfkp^R754W/R754W^and WT mice at different embryonic days. The statistically significant difference in left ventricular wall thickness between Pfkp^R754W/R754W^and WT mice in E15.5 and E17.5, whereas there was no statistical difference at E13.5. **Panel D** shows representative images of the immunofluorescence proliferation marker Ki67 in WT and Pfkp^R754W/R754W^ mice at E13.5, E15.5, and E17.5. **Panel E** shows the quantification of Ki67 levels in heart tissues of WT and Pfkp^R754W/R754W^ mice at different embryonic days. Cardiomyocyte proliferation was significantly lower in the mutant group compared to the normal control. **Panel F and G** show Ki67 heart immunofluorescence in WT and *Pfkp* cKO mice at E13.5, E15.5, and E17.5; cardiomyocyte proliferation was significantly lower in the *Pfkp* cKO group than in the normal control. Data in B, C, D, F, H, and I are represented as mean ± S.D, n=6 for each genotype per stage. Statistical significance of E15.5 in data B was determined with bonferroni test. Statistical significance of others data was determined with a two-tailed unpaired Student’s t-test. The scale bar indicates 30μm. (n.s., not significant (P>0.05) ; *P<0.05, **P<0.01, ***P<0.001).

### Cardiac Hyposplasia Manifested in *Pfkp* conditional knockout mice

To further clarify the function of PFKP during embryonic heart development, we generated a cKO mouse model with Myh6-driven Cre to eliminate Pfkp in cardiomyocytes. Myh6-Cre can specifically knock out target genes in mice myocardium from approximately embryonic day 10.5^21^. Subsequent assessment confirmed *Pfkp* expression across multiple organs and, importantly, revealed a significant reduction in PFK1 protein levels within the embryonic hearts of *Pfkp* cKO mice. The PFK1 enzyme activity was significantly reduced as well. While *Pfkp* cKO mice showed no significant changes in the LV and RV compact layer morphology at E13.5, pronounced thinning of the LV and RV compact layer was found at E15.5 and E17.5. And a smaller heart size was evident at E17.5. These findings strikingly paralleled the myocardial phenotypes observed in Pfkp^R754W/R754W^ mice, thereby conclusively establishing that Pfkp deficiency leads to abnormal myocardial development.

Given the capacity of Pfkp^R754W/R754W^ mice and *Pfkp* cKO mice to survive into adulthood, we generated separate survival curves for each group. The results showed that *Pfkp*^R754W/R754W^ mice and cKO mice generally exhibit survival rates consistent with normal life expectancy. In the cardiac pathological sections of 8-week-old mice, neither *Pfkp*^R754W/R754W^ mice nor *Pfkp* cKO mice exhibited any pathological manifestations of myocardial thinning. However, adult *Pfkp*^R754W/R754W^ and *Pfkp* cKO mice both exhibited reduced cardiac function. An adult cardiac compensatory mechanism might enable *Pfkp* functionally deficient mice to maintain normal cardiac morphology in adulthood.

### Reduced Cardiomyocytes Proliferation in PFKP Deficient Mouse Embryos and hPSC-CMs

Prior studies have demonstrated that Pfkp defects lead to a reduction in tumor cell proliferation, primarily due to impaired PFK1 enzymatic activity^22-24^. To investigate the necessity of Pfkp in embryonic cardiomyocyte proliferation, we examined myocardial proliferation in Pfkp^R75W/R754W^ and *Pfkp* cKO mice. Ki67 staining of heart tissue at E13.5, E15.5, and E17.5 revealed a significant decrease in cardiomyocyte proliferation in Pfkp^R75W/R754W^ mice (Fig.2d and 2e). Similarly, *Pfkp* cKO mice displayed reduced embryonic cardiomyocyte proliferation (Fig.2f and 2g), indicating that PFKP is essential for embryonic cardiomyocyte proliferation.

To validate the impact of PFKP on proliferation function during human cardiomyocyte development, we generated hPSC-CMs using patient-derived *PFKP* ^R755W/+^ and *PFKP* knockout cells. In line with the *in vivo* results, both *PFKP* mutations and *PFKP* KO hPSC-CMs led to a significant reduction in PFK1 enzyme activity in cardiomyocytes . *PFKP* ^R755W/+^ hiPS-CMs exhibited decreased proliferation in early-stage cardiomyocytes (10Day, 15Day hiPS-CMs) compared to WT and *PFKP* ^Correct/+^ hiPS-CMs. However, there were no significant differences in proliferation between late-stage *PFKP* ^R755W/+^ cardiomyocytes (30-day hiPS-CMs) and WT or *PFKP* ^Correct/+^ hiPS-CMs. *PFKP* KO hESC-CMs also showed decreased proliferation in immature cardiomyocytes.

We transfected PFKP protein using lentivirus in *PFKP* KO hESCs, and the results demonstrated that PFKP overexpression effectively restored the early cardiomyocyte proliferation caused by PFKP deficiency. These findings confirm that PFKP deficiency leads to reduced cardiomyocyte proliferation function.

### Abnormal Glycolytic Function and Decreased Downstream Metabolites in PFKP-Deficient Cardiomyocytes

We further investigated the glycolytic function of immature cardiomyocytes with *PFKP* mutations and *PFKP* KO (15D hPSC-CMs) through Seahorse ECAR experiments. The results showed that compared to WT and PFKP Correct/+ hiPS-CM, *PFKP* R755W/+ hiPS-CM exhibited significantly reduced glycolytic function. Similarly, *PFKP* KO hESC-CMs also displayed decreased glycolytic function in immature cardiomyocytes, and overexpression of PFKP in *PFKP* KO hESC-CM effectively restored the reduced glycolytic function caused by PFKP deficiency. These results indicate that PFKP functional defects can lead to abnormal glycolytic function in cardiomyocytes.

Glycolysis primarily generates ATP and provides precursors of macromolecular compounds such as nucleotides, proteins, and phospholipids necessary for cell mitosis and proliferation^25, 26^. To further investigate the effect of PFKP knockdown on the anabolic state of cardiomyocytes, we performed targeted mass spectrometry on immature hESC-CMs (Day 15). Compared with WT, *PFKP* KO hESC-CMs had 11 differential metabolites. Ten down-regulated metabolites in *PFKP* KO hESC-CMs, including Fructose-1,6-bisphosphate (F-1,6-BP), IMP, AMP, and other purine nucleotide synthesis substrates, glutamine, serine, and other amino acids. The results suggested that the synthetic metabolism deficiency of the above substances is one of the mechanisms of PFKP deficiency leading to the proliferation defect in cardiomyocyte development.

To confirm the impact of decreased glycolytic function on immature cardiomyocyte proliferation, we conducted *in vitro* experiments by modulating glycolysis through varying glucose concentrations through varying glucose concentrations during the early stages of cardiomyocyte development. A proportional decrease in hiPS-CM cardiomyocytes proliferation was observed with diminishing glucose concentrations. Similarly, administration of the glycolytic inhibitor 2-Deoxyglucose (2DG) to 10-day WT hESC-CMs demonstrated a reduction in early cardiomyocyte proliferation. Further investigations involved the inhibition of glycolytic activity in embryonic mice through intraperitoneal injections of 2DG to pregnant mice at day E12.5. Results indicated that compared to WT mice at E17.5, the addition of 2DG resulted in decreased thickness of the left ventricular muscles in mice and reduced myocardial proliferation. These findings suggest that a decrease in glycolysis leads to reduced early cardiomyocyte proliferation.

### F-1,6-BP Rescues PFKP Deficiency Phenotypes

F-1,6-BP is an endogenous intermediate of the glycolytic pathway generated as a consequence of PFK1 enzymatic activity^26^. To examine whether the replenishment of F-1,6-BP can effectively mitigate the myocardial hypoplasia phenotype associated with PFKP knockout (KO), we introduced F-1,6-BP on the 6th day of hESC differentiation in both the WT and *PFKP* KO experimental groups, assessing cardiomyocytes proliferation on the 10th day of differentiation (Fig. 3a). We observed that the introduction of F-1,6-BP successfully ameliorated the reduction in cardiomyocyte proliferation induced by *PFKP* KO (Fig. 3b). This observation prompted us to assess the impact of F-1,6-BP embryonic heart development in a murine model. Fetal mice were injected with F-1,6-BP via umbilical vein on embryonic E12.5 and their hearts were harvested at E17.5 for comprehensive phenotypic assessment (Fig. 3c). The administration of F-1,6-BP in the early embryonic period effectively rescues the thinning of LV and RV compact layer (Fig. 3d and 3e) and restores myocardial proliferation in *Pfkp* cKO mice (Fig. 3f and 3g).

**Figure 3.**
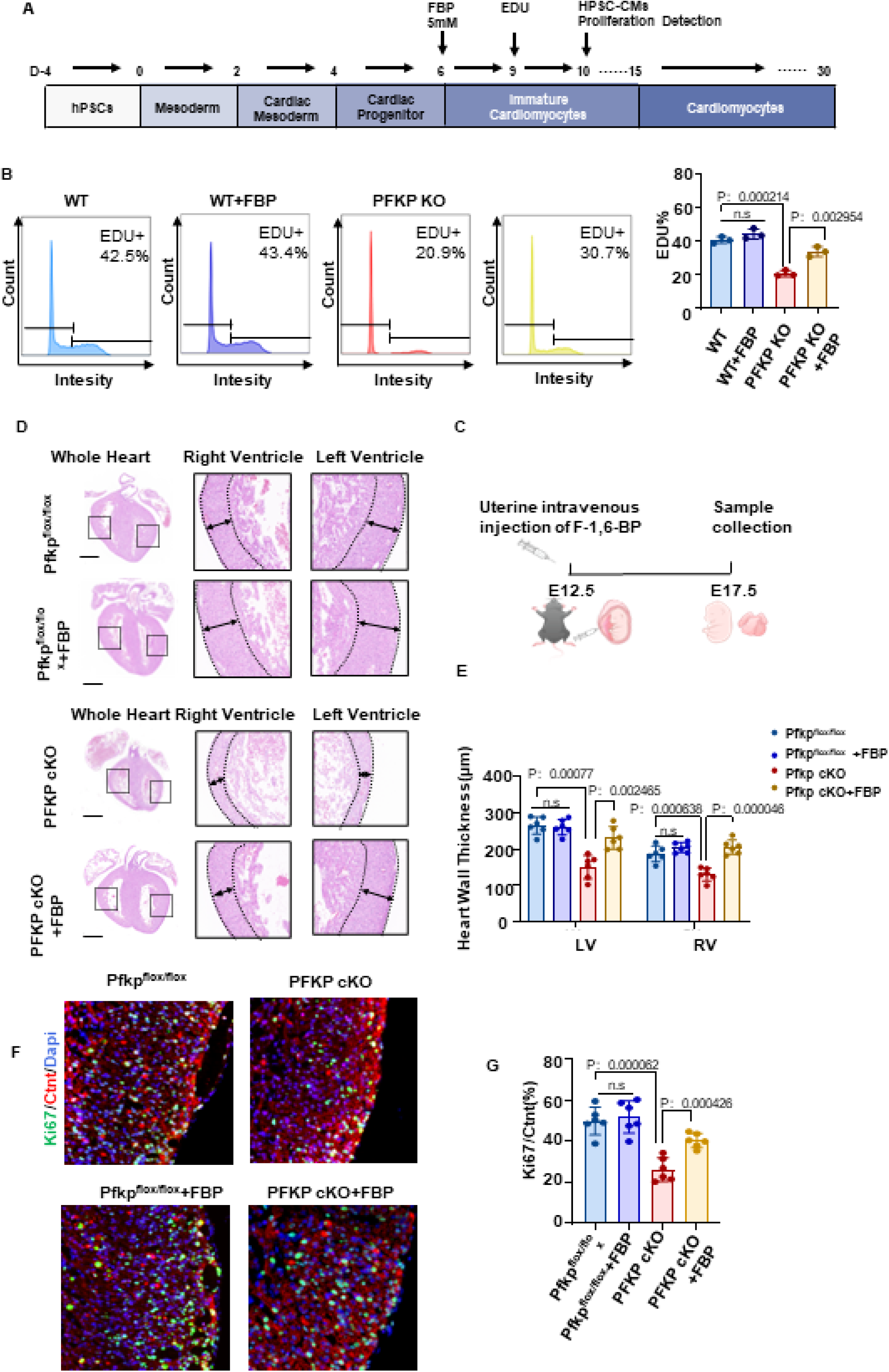
F-1,6-BP supplementation reverses PFKP deficiency-related myocardial hypoplasia. **Panel A** shows the flowchart for F-1-6-BP application to the *in vitro PFKP* KO hESC-CMs model. **Panel B** shows EdU coupling fluorescence flow cytometry and EDU positive percentage statistics of hESC-CM in WT, WT+ F-1,6-BP, *PFKP* KO, *PFKP* KO+ F-1,6-BP groups, PFKP deficiency leads to slower cardiomyocyte proliferation, which can be rescued by the supplementation of 5mM F-1,6-BP. **Panel C** shows a flowchart for treating pregnant *Pfkp* cKO mice with F-1,6-BP supplementation. 5mg F-1,6-BP dissolved in PBS for per pregnant mice via umbilical vein injection. **Panel D** shows a representative heart HE staining of WT+PBS, WT+ F-1-6-BP, *Pfkp* cKO+PBS, and *Pfkp* cKO+ F-1,6-BP at embryonic day 17.5. The scale bar indicates 600μm. **Panel E** shows a quantitative analysis of myocardial thickness in the above four groups, revealing that F-1-6-BP supplementation can rescue mice embryonic thinning of the left and right ventricular wall myocardium due to *Pfkp* knockout. (n=6 per group) **Panel F** shows representative immunofluorescence proliferation marker Ki67 images in WT+PBS, WT+ F-1,6-BP, *Pfkp* cKO+PBS, and *Pfkp* cKO+ F-1,6-BP at embryonic day 17.5. The scale bar indicates 30μm. **Panel G** shows the quantitative analysis of Ki67 immunofluorescence staining, revealing that F-1,6-BP supplementation can reverse decreased myocardial proliferation due to *Pfkp* knockout during the embryonic stage. (n=6 per group). Data in E and G are represented as mean ± S.D. Statistical significance was determined with a two-tailed unpaired Student’s t-test (n.s., not significant P>0.05 ).

### Preimplantation genetic testing and database screening

The above results indicate that *PFKP* R755W variant is the genetic cause of myocardial hypoplasia in family 1. Subsequently, the couple (Fig. 1A, III-1, and III-2) pursued assisted reproductive technology (Fig. 4a). Preimplantation genetic testing by MARSALA^20^ showed that the embryo was euploid (Fig. 4b) and did not carry the paternal *PFKP* R755W variant (Fig. 4c and 4d). After full consultation with the family and informed consent, the hormone replacement cycle - frozen embryo transfer is performed according to clinical routine. Ultrasound examination after embryo transfer confirmed the pregnancy. Regular prenatal check-ups revealed no abnormalities, particularly in the heart (Fig. 4e). The couple welcomed a healthy child, with the genetic variant and chromosome copy number confirmed via umbilical cord blood test, aligning with the embryo test results. No abnormalities were found in the neonatal physical examination and echocardiography at 80 days postpartum (data not shown).

**Figure 4.**
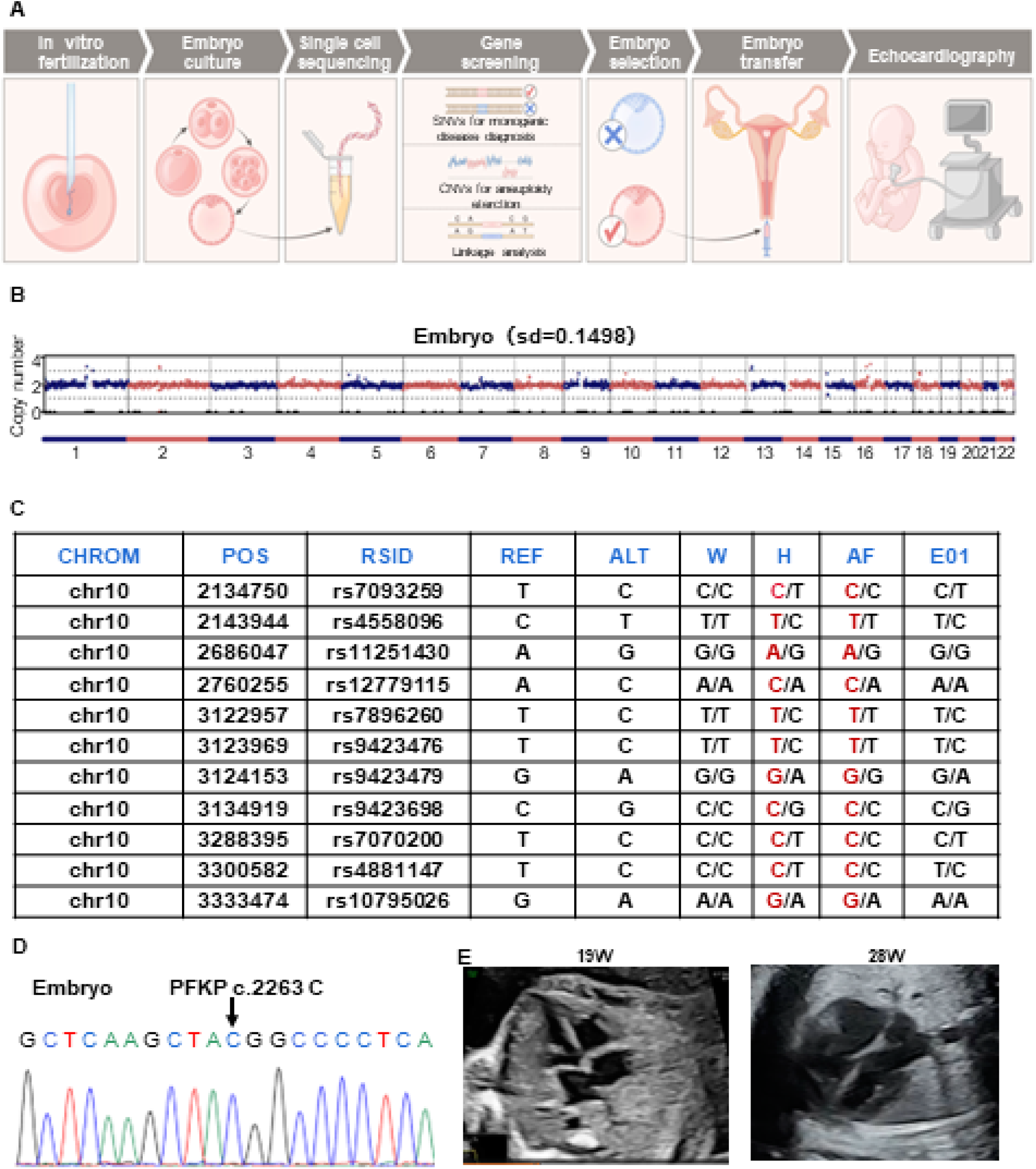
Assisted reproduction technology to get healthy offspring for the Family1. **Panel A** shows a flowchart for preimplantation genetic testing, intervention, and assisted reproductive conception. **Panel B** shows CNV analysis of the embryo (sex chromosomes not shown). **Panel C** Shows the Linkage analysis results of the case. The husband and the affected fetus carried variation sites in *PFKP*, while the wife and the embryo did not have *PFKP* pathogenic variant. Red letters indicated mutated alleles. CHROM, chromosome number; POS, SNP locations; REF, the SNPs of reference allele; ALT, the SNPs of alternative allele; W, wife; H, husband; AF, affected fetus; E, Embryo. **Panel D** shows Sanger sequencing results of the amplification products of the biopsy sample from the embryo. Results indicated the embryo was free of *PFKP* c.2263C>T. **Panel E** shows echocardiograms of IV-5 in the Family1 at prenatal 19, 28 weeks,which suggests normal heart structure.

In our quest to better understand the prevalence of PFKP defects, we conducted an initial screening of a local fetal heart disease exome database comprising 1300 fetal heart disease cases. Interestingly, we discovered the R755W mutation in yet another family within this dataset. Notably, the clinical presentation of patients from this second family closely mirrors that of family 1, and there is a clear co-segregation of the R755W mutation with the cardiac phenotype. The above results further improve the pathogenic evidence linking the *PFKP* R755W variation to myocardial hypoplasia.

## Discussion

Dysregulation of cardiomyocyte proliferation underlies the pathogenesis of the majority of CHD^13, 19, 27-29^. In model organisms like mice and zebrafish, impaired embryonic myocardial proliferation leads to postnatal cardiac morphological and functional impairments^30, 31^. Prior studies have also documented that proliferation abnormalities are associated with congenital cardiac malformations, including isolated ventricular myocardial hypoplasia^13, 19, 28, 29, 32^, both with and without structural defects. In this study, two familial clusters of myocardial dysplasia with or without structural malformations were detected by elaborate phenotypes through fetal echocardiography. The clinical manifestations of the fetal probands provide an opportunity to investigate embryonic cardiac pathogenesis. Apart from this, considering family carriers across the lifespan allows for the exploration of factors influencing phenotypic differences, especially the intrauterine environment. This study marks the first instance of verifying the pathogenic genes and mechanisms responsible for the disturbance in embryonic myocardial proliferation, which could pave the way for common mechanisms as well as strategies for prevention and intervention in CHDs.

Previously, PFKP function was mainly investigated in tumors^22-24^, with limited attention and reporting in the context of the embryonic heart. For the first time, we provided evidence that *in vivo* and *in vitro* PFKP deficiency disrupts cardiomyocyte proliferation by impacting glycolysis, consequently resulting in myocardial hypoplasia. Furthermore, in the human context, we used preimplantation genetic testing for monogenic disorders to prevent the transmission of PFKP pathogenic variants, resulting in the development of normal-hearted, healthy fetuses. Interestingly, in 8-week-old adult mice, our study demonstrated that *Pfkp* cKO hearts did not show structural changes but exhibited a reduction in systolic function. The relatively mild phenotype observed in adult *Pfkp* cKO mice suggests that past intrauterine environment, postnatal alterations in gene expression dominance, and compensation by isozymes are likely the primary mechanistic factors underlying this outcome. These findings serve as the basis for our clinical interventions such as prenatal counseling and preconception genetic blocking. Glycolysis is the primary energy supply for the developing embryonic heart^18, 30, 33, 34^, and its deficiency is recognized as a primary cause of cardiac development disorder^31, 33-36^. Recent research has also highlighted the co-regulation of metabolic pathways, including anaerobic glycolysis, as a contributing factor to myocardial proliferation and regeneration following cardiac injury^30, 36, 37^. Consistent with our original hypothesis, impaired glycolytic function significantly curtails the proliferative capacity of cardiomyocytes, culminating in a myocardial hypoplasia phenotype. Importantly, the administration of F-1,6-BP, the processed substrate of PFK1, effectively mitigated the manifestation of myocardial hypoplasia in *Pfkp* cKO mice. Hence, investigating glycolytic deficiencies might present potential as a viable strategy for CHD in utero interventions.

Consequently, the systematic identification and screening of pathogenic variants within glycolysis-related genes emerge as indispensable tools in advancing the realms of CHD prevention and intervention, alongside the management of other fetal heart disorders. Our future endeavors will focus on more investigations of fetal heart disease phenotypes associated with PFKP pathogenic variants, thereby strengthening the foundations for comprehensive fetal CHD screening. Furthermore, a much deeper understanding of the functions of PFKP in myocardial proliferation will unveil additional therapeutic targets, extending the scope of potential interventions for myocardial hypoplasia.

### Author contributions

Conceptualization: S.Y.Z.,H.R.S.,X.Y.H and R.M.L; Methodology: S.Y.Z.,H.R.S., X.Z.,X.W.L and L.Y.Y.; Resources: Y.Z., X.Y.G., J.C.H. ,T.Y.,H.K.L and J.Q.F.; Writing–Original Draft, S.Y.Z.,H.R.S. and R.M.L; Writing –Review & Editing, Y.H.H, F.L.,J.Q and H.J.Z; Project Administration: X.Y.H.; Funding Acquisition: Y.H.H. and X.Y.H.

## Data Availability

All data produced in the present work are contained in the manuscript

## Acknowledgments

Many thanks to Professor Dong Zhao of the Department of Epidemiology, Beijing Anzhen Hospital, Capital Medical University, Beijing Institute of Heart, Lung, and Blood Vessel Diseases. We like to thank Professor Yimin Hua and Yifei Li of the Department of Pediatrics, West China Second University Hospital, Sichuan University. This study was sponsored by the National Natural Science Foundation of China (No U21A20523 and 82170301 to Yihua He; No. 82200334 to Xiaoyan Hao; No. 82100322 to Tong Yi).

