## Supplemental Methods for "Prenatal glycolysis restoration can rescue myocardial hypoplasia caused by platelet isoform of phosphofructokinase 1(PFKP) deficiency": Preprint supplementary appdendix1.pdf

### Genetic analysis

Whole genome sequencing (WGS) was performed with the DNA of patients and controls. Then, linkage analysis and variant calling, annotation, filtering, and prioritization were performed. The pathogenicity of sequence variants was assessed according to ACMG guidelines. Full details are provided in the the TableS1.

Supplementary Appendix

### Genome sequencing AND variants calling

gDNA was extracted from whole blood or umbilical cord using the Qiagen DNA Blood Midi/Mini kit (Qiagen GmbH, Hilden, Germany). DNA libraries were prepared using the TruSeq PCR-free DNA HT sample preparation kit (Illumina, San Diego, CA USA) according to the manufacturer's protocol. The size distribution and concentration of the libraries were respectively determined by Agilent 2100 Bioanalyzer and qualified using real-time PCR. DNA library was sequenced on Illumina Novaseq 6000 platform for paired-end 150bp reads. Then, raw image files were processed using Bcl To Fastq (Illumina) for base calling and generating raw data. Low-quality sequencing reads were filtered out using a quality score 20 (Q20). The reads were aligned to the NCBI human reference genome (hg19/GRCh37) using the BWA. BAM files were subjected to single nucleotide polymorphism (SNP) analysis, duplication marking, indel realignment and recalibration using GATK[1].

GATK[1], expansionHunter[2], CNVnator[3], and LUMPY[4] were used to detect the per-sample SNV/INDEL, STR,CNV and SV. Pedigree SNV/INDEL spectrum was detected by GATK. SNV/INDEL low-depth (per-sample depth < 6), low variant quality (<100), low map quality (<55), or low genotype quality (per-sample quality < 30) were removed. Per-sample STRs with low-depth (depth < 6), low variant quality (<100) were removed. Pedigree STRs were merged by bcftools[5]. Per-sample CNVs with Q-value > 0.05, fraction of reads with zero map quality > 0.5, or fraction of gap > 0 were removed. Pedigree CNVs coordinates with 1 bp overlap were merged together by bedtools[6]. Pedigree SVs were genotyped SVTyper[7]. SVs with any of the quality control criticisms employed from Abel's study[8] were excluded: (1) The proportion of split-read and paired-end read counts < 10%; (2) mean sample quality < 150; (3) the deletion size < the insert size of sequencing library estimated by SVTyper; (4) deletion copy-number estimated by CNVnator3 > 0.5 or duplication copy-number < 1.5.

**Linkage analysis**

According to the disease status in pedigree and incidence rate, we assumed this is an autosomal dominant disease caused by rare mutation. Rare bi-allele SNVs were used for linkage analysis. Allele frequency was annotated by VEP with gnomAD[9] and ChinaMAP[10] database. A threshold of allele frequency < 0.005 in gnomAD-EAS and ChinaMAP was used to determine the rare mutation marker. Merlin[11] was used to estimate the linkage association of marker and disease. Firstly, single-point logarithm of the odds (LOD) scores was calculated for all markers from whole genome. Subsequently, an expected LOD (ELOD) score for this pedigree assuming a dominant model was calculated by SwiftLink. A threshold of single-point LOD score ≥ ELOD score was used to determine the peak of linkage association. Finally, multi-point analysis of linkage peak was used to determine the linkage region of genome. Co-segmentations were identified from the rare SNVs, INDELs, STRs, SVs, and CNVs in the region. Statistical evidence for the pathogenicity of co-segregated SNV was indicated by full-likelihood Bayes factor[12] (FLB) that calculated by segregatr R-package.

**Sequence variant filtering and prioritizing**

Sequencing variant filtering were performing as previously described[13]. Analysis was performed on all sequenced genomic regions including both exonic and intronic where good data coverage was guaranteed. Variant frequencies were determined in dbSNP147 (https://www.ncbi.nlm.nih.gov/SNP/), the 1000 Genomes Project(http://www.internationalgenome.org/),Exome Variant Server (http://evs.gs.washington.edu/EVS), ExAC (http://exac.broadinstitute.org/), gnomAD[9], ChinaMAP[10] and in-house database to remove common SNPs (minor allele frequency> 0.1%). SIFT (http://sift.jcvi.org), PolyPhen-2 (http://genetics.bwh.harvard.edu/pph2), MutationTaster(http://www.mutationtaster.org),REVEL(https://sites.google.com/site/revelgenomics/) and CADD (http://cadd.gs.washington.edu) were used for predicting the pathogenicity of missense variants, while Human Splicing Finder (http://www.umd.be/HSF) and

MaxEntScan (http:// genes.mit.edu/burgelab/maxent/Xmaxentscan_scoreseq.html) were used for evaluating the effects on splicing. Moreover, databases such as OMIM(http://omim.org/), ClinVar(http://www.ncbi.nlm.nih.gov/clinvar), and Human Gene Mutation Database(http://www.hgmd.org) were used to determine variant harmfulness and pathogenicity where appropriate. Finally, filtering and prioritizing were performed to investigate potential detrimental variants such as nonsense, missense, frameshift, indel, and variants impacting splice for variant interpretation and classification according to ACMG guidelines[14].

**IVF-ICSI and embryo biopsy**

After informed consent, the couple went through a controlled ovarian hyperstimulation cycle and got 7 mature metaphase Ⅱ oocytes retrieved, which were then fertilized through intracytoplasmic sperm injection (ICSI). Fertilization was confirmed 17 hours later, with 2PN observed in 5 zygotes. The zygotes were then cultured in G1-plus media (Vitrolife, Göteborg, Sweden) in a 37°C, 6% CO2 incubator (Thermo Scientific, Waltham, MA, USA). Five embryos successfully developed to Day 3 and got transferred to G2-plus media (Vitrolife, Göteborg, Sweden). Trophectoderm biopsy was performed on only 1 blastocyst which developed on Day 6. A sample of trophoblasts was collected for the later whole genome amplification (WGA) process. The blastocyst was then cryopreserved by vitrification.

**WGA and sequencing**

WGA of the biopsied sample was carried out with multiple annealing and looping-based amplification cycles (MALBAC). Single-Cell WGA Kit (Yikon Genomics Inc., China). WGA product was then prepared for the mutated allele revealed by sequencing with aneuploidy and linkage analyses (MARSALA), which enables the simultaneous detection of aneuploidy, known point variation, and linkage analyses in a single procedure, previously described by (Live births after simultaneous avoidance of monogenic diseases and chromosome abnormality by next-generation sequencing with linkage analyses.

**Mice transgenic lines**

We used CRISPR/Cas9 technology to introduce the target point mutation into C57BL/6J mice's zygotes through homologous recombination repair. Cas9 mRNA and guide RNA (gRNA) were synthesized via in vitro transcription, and the oligo donor DNA was synthesized. Subsequently, we microinjected Cas9 mRNA, gRNA, and donor DNA into C57BL/6J mice's zygotes to generate the F0 generation of mice. The F0 Pfkp R755W/+ males were bred to wild-type mice for two generations, and the genotypes of Pfkp R754W/+ founding males and all F2 offspring were confirmed by PCR and Sanger sequencing. F2 mice (Pfkp R754W/+) were intercrossed, and hearts derived from homozygous, heterozygous, and wild-type littermates were collected at desired developmental stages for specific assays, the breeding strategy is consistent across all assays unless where is specified. The details of gRNA, primers, and oligo donor sequences are provided in Table S2.

We generated a cardiac-specific Pfkp knockout (Pfkp-CKO) mouse line by crossing Pfkp flox/flox mice with Myh6 Cre (Cre+/−) mice. All mouse strains used were of the C57/B6 background. For experiments, embryos and neonatal mice of both sexes were grouped according to their genotypes. The day of detecting vaginal plugs at noontime was designated as E0.5, while the morning of observing newborns was designated as P0. Genotyping was performed using PCR on yolk sac or tail samples from the mice, utilizing Cre and allele-specific primers (Table S2).

**Cell culture and cardiac differentiation**

H9 and hiPS cells were maintained on feeder-free Matrigel (Corning) and fed daily with E8 medium (Cellapy). Cells were routinely passaged every 3 days at 70–80% confluency using 0.5 mM EDTA in PBS without $MgCl_2$ or $CaCl_2$ (HyClone, USA). The cells were cultured at 37 °C with 5% $CO_2$. HPSCs were differentiated into hPSC-CMs using a small molecule-based method as previously described[15]. hPSC-CMs were purified with the lactate metabolic selection method[16].

**Genome editing**

We designed Single-guide RNAs (sgRNAs) to target exon 4 of the PFKP genome using online tools available at https://design.synthego.com/. Among the two sgRNAs tested (TCCCAGGGCGGGACGATCAT and GTCCCAGGCCTTCCGCACGC), the second sgRNA demonstrated higher editing efficiency. This second sgRNA was selected for genome editing in H9 cells. To facilitate this, we cloned the sgRNA into the epiCRISPR plasmid, following a previous publication[17]. We dissociated 2 × $10^6$ H9 cells using 0.5 mmol/L EDTA and subsequently electroporated these cells with 3 μg of the epiCRISPR plasmid in a solution composed of 82 μL of stem cell P3 solution and 18 μL of supplement (Lonza). The electroporation procedure was carried out using the 4D nucleofector system with the CA137 program (Lonza). After transfection, the cells were plated onto Matrigel-coated 10cm Petri dishes and cultured in E8 medium supplemented with 10 µM Rho kinase inhibitor Y-27632 (MedChemExpress) for the initial 24 hours. On day 3, the cells underwent puromycin selection at a concentration of 0.3 µg/mL.

**Reprogramming of peripheral blood mononuclear cells (PBMC) and homologous recombination repairing of PFKP R755W hiPS cell line**

Peripheral blood mononuclear cells (PBMCs) of the patient and normal people were isolated within 2 hours of collection using Vacutainer® CPT™ Cell Preparation Tubes with Sodium Heparin (BD Biosciences) and separated by centrifugation at 1800 relative centrifugal force (rcf) for 30 minutes at room temperature (RT). The freshly isolated PBMCs were then seeded at a density of $0.6\times 10^6$ cells in Expansion Medium (EM). Nine days later (Day 0), the cells were transduced using Sendai virus delivery with the CytoTune®-iPS 2.0 Sendai Reprogramming Kit (Thermo Fisher Scientific), following the manufacturer's instructions. The cell-containing plate was then centrifuged at 900 rcf for 90 minutes at RT and incubated for 24 hours. The following day, the cells were collected, centrifuged at 300 rcf for 5 minutes at RT, and seeded in fresh EM (Day 1). Two days later (Day 3), the cells were transferred to a 6 cm dish coated with 0.1% gelatin and co-cultured with $0.8 \times 10^6$ Mitomycin C-treated mouse embryonic fibroblasts (MEF) in QBSF-60 medium (Quality Biological) supplemented with 50 µg/ml ascorbic acid. Seven days post-transduction (Day 7), the medium was replaced with mTESR-1 medium (Stem Cell Technologies). Colonies displaying an embryonic stem (ES)-like appearance were manually isolated based on morphology between Day 21 and Day 27 post-transduction and subsequently cultured as induced pluripotent stem cells (iPSCs).

Patient-derived hiPSC cells were transfected with sgRNA/Cas9 RNP + ssODN complexes (for sequence details, please refer to the Supplementary Appendix). Subsequently, a portion of the transfected cells was harvested for sequencing. Mixed cell populations were amplified, individual clones were selected, and PCR amplification, as well as first-generation sequencing identification, were carried out. Human iPSC cultures were maintained on plates coated with Matrigel (Corning) in mTESR-1 medium (Stem Cell Technologies), following the manufacturer's instructions. All cells were cultured at 37°C in a humidified atmosphere containing 5% CO2.

**Lentivirus transfected cells**

**D**NA sequences encoding isoforms of human PFKP WT and PFKP R755W were PCR-amplified from a human NCBI library and cloned into pcDNA6/myc-His (Invitrogen) using NEBuilder (NEB) to construct pcDNA6-PFKPWT-myc-Flag, pcDNA6-PFKPR755W-myc-Flag. The plasmids were verified by sequencing at the Support Unit for Bio-Material Analysis of RIKEN RRD. 24 hours before lentiviral transfection, cardiomyocytes were seeded in a 12-well plate at a density of $4×10^5$ cells per well, while HEK293T cells were plated at a density of $2×10^5$ cells per well. The cell density during lentiviral transfection remained at approximately $4×10^5$ cells per well. The viral suspension was diluted to achieve an MOI of 5 and incubated at 37°C for 24 hours. Subsequently, the virus-containing medium was replaced with a fresh medium. After an additional 72 hours of incubation, pronounced fluorescent expression became evident.

**Histology**

For histological studies, hearts were harvested at the indicated time points. The entire hearts were fixed overnight at 4 °C in 4% paraformaldehyde (PFA, Sigma). Subsequently, the hearts underwent dehydration through a series of increasing ethanol concentrations and were then embedded in paraffin. The embryonic mice hearts were longitudinally sectioned 3μm for each tissue section. Hematoxylin and eosin staining were performed following established protocols (Heallen, T. et al. Hippo signaling impedes adult heart regeneration. Development 140, 4683–4690, 2013). The HE-stained tissue sections were scanned using the Pannoramic MIDI II slide scanner and analyzed using the caseviewer software.

**Echocardiography**

The cardiac function of 18-week-old mice was evaluated through a series of echocardiographic examinations. Left ventricular systolic function was assessed using echocardiography (Visual Sonics Vevo 2100) with the mice under 1.5% isoflurane anesthesia while maintaining spontaneous ventilation. Two-dimensional B-mode imaging was employed to obtain long-axis projections, along with guided M-mode images. The calculation of left ventricular ejection fraction (LVEF) and fractional shortening (FS) was performed based on measurements of end-diastolic and end-systolic dimensions obtained from the M-mode ultrasound scans.

**Immunofluorescence**

Before the cells were tested, the normal culture medium was replaced with a medium containing 10 μM EdU ( C0071S, Beyotime) 24 h. Cells were fixed with 4% paraformaldehyde (PFA). Hpsc-CMs were permeabilized with 1.0% Triton X-100 (Sigma) for 15 minutes. Then the cells were subjected to a blocking step using 3% BSA (Sigma) at room temperature for 30 minutes. Prepare the click reaction solution and incubate it in the dark for 30 minutes at room temperature. Wash slides three times with 3% BSA for five minutes each time.

The cells were initially treated by fixation in a solution containing 4% paraformaldehyde (PFA), followed by permeabilization with 1.0% Triton X-100 (Sigma) for 15 minutes. Subsequently, they were subjected to a blocking step using 3% BSA (Sigma) at room temperature for 30 minutes. Next, the cells were incubated with the primary antibody overnight at 4°C and later incubated with a secondary antibody at 37℃ for 1 hour. Following this, the cells were subjected to three 5-minute washes with PBST. Finally, they were counterstained with DAPI for 5 minutes, and the images were acquired using a Leica DMI 4000B confocal microscope (Leica).

The paraffin-embedded heart slides were de-paraffinized with xylene and then rehydrated in decreasing concentrations of ethanol (100, 100, 95, 90, and 80%) and water. The heart slides underwent antigen retrieval by microwaving in citrate solution for 10 min; then, the slides were blocked with 10% goat serum (Invitrogen) and incubated with the primary antibody overnight at 4 °C. The next day, the slides were washed in PBST three times and incubated with the appropriate fluorescent secondary antibody at 37℃ for 1 hour. The slides were washed in PBST three times and then stained with DAPI to label the nuclei. The specific antibodies used are listed in Table S2. The results were analyzed using the ImageJ software program.

**Western blot**

Select 50 mg of human and mouse ventricular tissue to extract protein, and put them into 200 μl of pre-configured protein lysis solution (containing Mammalian Protein Extraction Reagent (Thermo, #78501, USA) supplemented with 5 mM EDTA (Thermo, #1861275), along with a protease inhibitor cocktail (Thermo, #1861278) and a phosphatase inhibitor cocktail (Thermo, #1862495)). The cells were washed with cold PBS and resuspended in protein lysis solution. The samples were placed on ice for 30 minutes, with intermittent agitation every 10 minutes, followed by centrifugation at 12,000 rpm for 15 minutes. The protein concentration in the resulting supernatant was determined using the BCA method. Equivalent amounts of denatured protein were separated by electrophoresis on 8–12% sodium dodecyl sulfate-polyacrylamide gels, depending on the protein size. The gel was subsequently transferred to a PVDF membrane at 300 mA for 90 minutes using a gel transfer apparatus (Bio-Rad). Afterward, the membrane was blocked with 5% nonfat milk powder at room temperature for 1 hour. Primary and secondary antibodies, with appropriate concentrations as listed in Table S2, were used for membrane incubation. All primary antibodies were incubated at 4°C overnight, while secondary antibodies were incubated at room temperature for 1 hour.

**Quantitative reverse transcription PCR (qRT-PCR).**

Quantitative reverse transcription PCR (qRT-PCR) was conducted as follows: Total RNAs were extracted from ventricles or cultured cardiomyocytes (CMs) using TRIzol reagent (Life Technologies). Subsequently,1μg RNA was reverse transcribed into cDNA by the PrimeScript™ reverse transcription system (Takara)according to the manufacturer's instructions. qRT-PCR was then performed on iCycler iQ5 (Bio-Rad) with 2 × SYBR Master Mix (Takara). Specific primer sequences are provided in Table S2. To normalize the relative expression of each gene, 18sRNA was used as an internal reference, and calculations were performed utilizing the 2−ΔΔCT method. For robustness, biological replicates were carried out using three individual samples of each genotype, and technical triplicates were included for each qPCR run.

**PFK enzymatic assay.**

We assessed the activity of the PFK1 enzyme in cells and tissues using the 6-Phosphofructokinase Activity Assay Kit (Abcam, ab155898). Fresh embryonic day 17.5 mouse heart tissue was harvested, and the requisite number of cells (initially recommended: 2 x $10^6$ cells) was collected for each assay. Protein quantification in the samples was performed, with an initial recommended protein input of 5μg per sample. Subsequently, we followed the manufacturer's instructions provided with the kit. To determine PFK1 enzyme activity, we generated enzyme activity curves by measuring absorbance at 450nm using a microplate reader. The time during which the curve displayed a stable slope was selected for the calculation of enzyme activity. The control group was set as 100% and statistical analyses were conducted. Time-absorbance curves were plotted using Igor software, and statistical graphs were generated using Prism 8 (GraphPad).

**Seahorse ECAR measurement**

For the Seahorse XF ECAR assay, 5×104 hPSC-CMs/well were seeded onto an XFe24 cell culture microplate coated with Matrigel 48 hours before the assay. To prepare the glycolysis assay media, we prepared a solution of 2mmol/L glutamine (103579-100, Agilent) using Seahorse XF Basal Medium (103334-100, Agilent). Approximately 60 minutes before the assay, we washed the cells with 1 ml of glycolysis assay media and incubated them for 45 minutes at 37 °C without CO2. Afterward, we added 450 μL of detection solution to each well. The assay cartridge was loaded with XF Cell Glycolytic Stress Test compounds (10 µM glucose, 1 μM oligo, 50 mM 2-DG). The XF Cell Culture Microplate was promptly inserted into the Seahorse XFe Analyzer, and the XF Cell Mito Stress Test was initiated. Following the measurement of extracellular acidification rate (ECAR), hPSC-CMs were dissociated, and the cell count in each well was determined. The ECAR values were normalized per 10,000 cells. Data analysis was performed using Wave software (Agilent).

**Flow cytometry**

Before cell testing, the culture medium was substituted with a medium containing 10 µM EdU (C0071S, Beyotime) for 24 hours. Hpsc-CMs were isolated using CardioEasy CM dissociation buffer I (Cellapy) at 37℃ for 15 minutes and CardioEasy CM dissociation buffer II (Cellapy) at 37℃ for 30 minutes. Following this, the cells underwent three PBS washes and were fixed with 4% paraformaldehyde (PFA, Sigma) for 30 minutes at room temperature. After three 3% BSA washes, The cells were permeabilized with 1.0% Triton X-100 (Sigma) for 15 minutes. After another three 3% BSA washes, the cells were treated with the click reaction solution for 30 minutes at room temperature. Subsequently, the samples were rinsed with PBS and assessed using FACS analysis (EPICS XL). After the PBS wash, the cells were resuspended following centrifugation in PBS and stored on ice until flow cytometry was conducted. The results were analyzed using the FlowJo X software program.

### Metabolomic analysis

Unsupervised PCA (principal component analysis) was performed by statistics function prcomp within R (www.r-project.org). The data was unit variance scaled before unsupervised PCA. The HCA (hierarchical cluster analysis) results of samples and metabolites were presented as heatmaps with dendrograms, while Pearson correlation coefficients (PCC) between samples were calculated by the cor function in R and presented as only heatmaps. Both HCA and PCC were carried out by R package pheatmap. For HCA, normalized signal intensities of metabolites (unit variance scaling) are visualized as a color spectrum. Significantly regulated metabolites between groups were determined by absolute LogFC (fold change).
